# Non-Culprit Left Main Coronary Artery Disease in Acute Myocardial Infarction Complicated by Cardiogenic Shock

**DOI:** 10.1101/2022.10.13.22281047

**Authors:** Ik Hyun Park, Woo Jin Jang, Ju Hyeon Oh, Jeong Hoon Yang, Young Bin Song, Joo-Yong Hahn, Seung-Hyuk Choi, Hyeon-Cheol Gwon, Chul-Min Ahn, Cheol Woong Yu, Hyun-Joong Kim, Jang-Whan Bae, Sung Uk Kwon, Hyun-Jong Lee, Wang Soo Lee, Jin-Ok Jeong, Sang-Don Park

## Abstract

**Objectives:** We evaluated the clinical impact of residual non-culprit left main coronary artery disease (LMCAD) on prognosis in patients undergoing emergent percutaneous coronary intervention (PCI) for acute myocardial infarction (AMI) complicated by cardiogenic shock (CS).

**Methods:** A total of 429 patients who underwent PCI for AMI complicated by CS was enrolled from 12 centers in the Republic of Korea. The patients were divided into two groups according to presence of non-culprit LMCAD or not: the LMCAD non-culprit group (n = 43) and the no LMCAD group (n = 386). Primary outcome was major adverse cardiac event (MACE, defined as a composite of cardiac death, myocardial infarction, or repeat revascularization). Propensity score matching analysis was performed to reduce selection bias and potential confounding factors.

**Results:** During a 12-month follow-up, a total of 168 MACEs occurred (LMCAD non-culprit group, 17 [39.5%] vs. no LMCAD group, 151 [39.1%]). Multivariate analysis revealed no significant difference in the incidence of MACE at 12 months between the LMCAD non-culprit and no LMCAD groups (adjusted hazard ratio [HR] 0.94, 95% confidence interval [CI] 0.56 to 1.58, *p* = 0.817). After propensity score matching, the incidence of MACE was still similar between the two groups(HR 1.07; 95% CI 0.49 to 2.36; *p* = 0.857). The similarity of MACEs between the two groups was consistent across a variety of subgroups.

**Conclusions:** After adjusting for baseline differences, residual non-culprit LMCAD does not appear to increase the risk of MACEs at 12 months in patients undergoing emergent PCI for AMI complicated by CS.

## INTRODUCTION

Left main coronary artery disease (LMCAD) is incidentally identified in 5<7% of patients undergoing coronary angiography [1]. Culprit LMCAD in patients with acute myocardial infarction (AMI) is considered a high-acuity and critical status because cardiogenic shock (CS) or cardiac arrest is a frequent complication associated with higher mortality. The question then arises as to whether non-culprit LMCAD is also related to adverse clinical outcomes in AMI complicated by CS. Do we have to treat emergently the non-culprit LMCAD for better outcomes in AMI patients with CS? The COMPLETE trial showed that complete revascularization (CR) including non-culprit coronary stenoses, either at the time of the index procedure or as a staged procedure, is superior to a culprit-only strategy in reducing the cardiovascular risk among AMI patients with multi-vessel disease [2]. The ISCHEMIA study reported that conservative medical treatment had similar mortality compared to an initial invasive strategy for stable coronary disease including multi-vessel disease and LMCAD [3]. These two previous studies did not include AMI patients complicated by CS and we still have no data about the prognostic effect and optimal treatment strategy of non-culprit LMCAD in AMI patients with CS. This study evaluated the clinical impact of residual non-culprit LMCAD on long-term clinical outcomes in patients undergoing percutaneous coronary intervention (PCI) for AMI complicated by CS.

## METHODS

### Study Population

The design of the RESCUE (REtrospective and prospective observational Study to investigate Clinical oUtcomes and Efficacy of left ventricular assist device for Korean patients with cardiogenic shock, NCT02985008) registry has been described previously [4]. In brief, between January 2014 and December 2018, a total of 1,247 CS patients older than 19 years was recruited from 12 Korean tertiary care centers. The criteria for CS included systolic blood pressure <90 mmHg for 30 minutes or need for inotrope or vasopressor support to achieve a systolic blood pressure >90 mmHg, and the presence of pulmonary congestion and signs of impaired organ perfusion (altered mental status, cold skin, urine output <0.5 mL/kg/h for the previous six hours, or blood lactate >2.0 mmol/L). Patients with out-of-hospital cardiac arrest, other causes of shock, and those who refused active treatment were excluded from this registry.

Among the 836 patients who presented with CS caused by AMI, data from 695 patients who underwent PCI were included in the final analysis. Reasons for additional exclusions were: 26 patients for whom coronary angiography was not attempted, 38 patients who did not receive revascularization or who failed culprit lesion PCI, 28 patients who did not have images of coronary angiography, 42 patients who underwent coronary artery bypass grafting, and 7 patients with vasospasm. For this study, we further excluded 256 patients who had only a culprit lesion of AMI or an identified culprit LMCAD and 10 patients for whom culprit lesion or vessel information was unavailable. Data from the remaining 429 patients were evaluated, and subjects were divided into 2 groups according to the presence or absence of residual non-culprit LMCAD after emergent PCI for culprit lesions (**Figure 1**).

**Figure 1.**
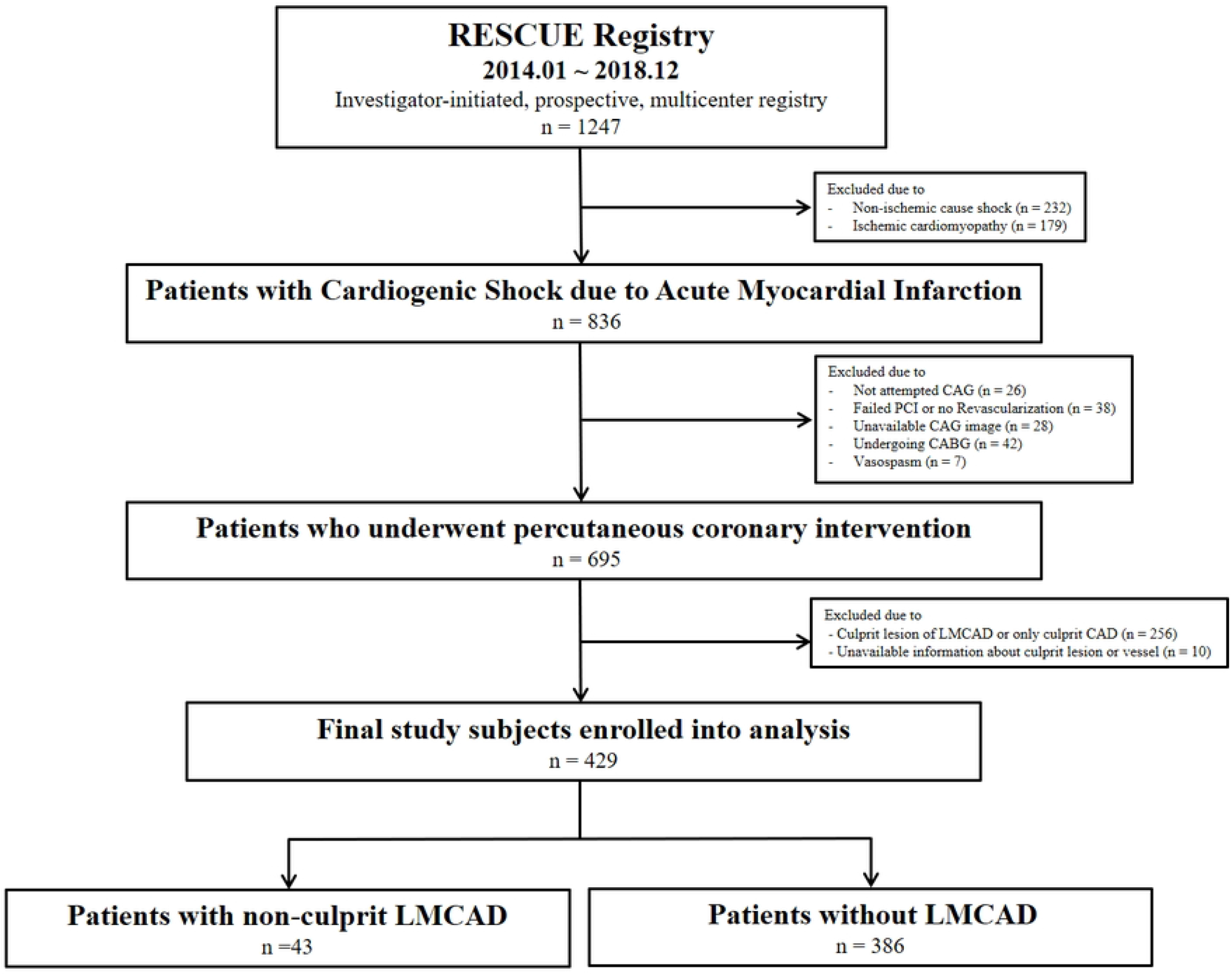
Schematic illustration of study cohort selection. CABG = coronary artery bypass grafting; CAD = coronary artery disease; CAG = coronary angiography; LMCAD = left main coronary artery disease; PCI = percutaneous coronary intervention

### Data Collection

Clinical patient demographics, in-hospital management, laboratory data, procedural data, and outcome data were collected by independent clinical research coordinators using web-based case report forms. All baseline data were measured on admission of patients. Additional information was obtained from medical records or telephone contact, if necessary. Institutional review board approval was obtained at each of the participating sites. The institutional review boards of the participating centers waived the requirement for informed consent in retrospectively enrolled patients, and informed consent was obtained before enrollment in all prospectively enrolled patients.

### PCI and Pharmacologic Therapy

PCI was performed according to standard techniques [5]. Unfractionated heparin or low molecular-weight heparin was used for anticoagulation during the procedure. The decision to perform thrombus aspiration, pre-dilation or post-dilation, or to use glycoprotein IIb/IIIa inhibitors was left to the operator. The length and diameter of stents were not restricted. The use of intravascular imaging or fractional flow reserve was performed at the operator’s discretion. All patients who were not taking aspirin or a P2Y12 inhibitor received a loading dose of aspirin (300 mg) or P2Y12 inhibitor (clopidogrel 300 - 600 mg, ticagrelor 180 mg, or prasugrel 60 mg). After the procedure, aspirin (100 mg orally once daily) was used indefinitely; clopidogrel (75 mg orally once daily), ticagrelor (90 mg orally twice daily), or prasugrel (10 mg orally once daily) was maintained. Anticoagulation during PCI was performed using low-molecular-weight heparin or unfractionated heparin to achieve an activated clotting time of 250 to 300 seconds. All patients were recommended to receive optimal pharmacological therapy, including statins, beta-blockers, or renin-angiotensin system blockade if indicated; the responsible clinicians determined the duration of dual antiplatelet therapy [6,7].

### Study Outcomes and Definitions

The primary outcome of this study was major adverse cardiac event (MACE), defined as a composite of cardiac death, myocardial infarction, or repeat revascularization. Secondary outcomes were consistent with the individual components of the primary outcome, as well as all-cause death, and re-hospitalization due to heart failure. Clinical events were defined based on recommendations from the Academic Research Consortium [8]. Analyses were truncated at 12 months of follow-up due to the different follow-up durations.

### Statistical Analysis

Categorical variables are presented as count and percentage and were compared using the χ^2^ test or Fisher’s exact test as appropriate. Continuous variables are presented as mean ± standard deviation or as median (25th percentile to 75th percentile) for variables lacking a normal distribution. Analysis of continuous variables was performed using Student’s t-test or Wilcoxon rank-sum test. Survival curves were generated using Kaplan–Meier estimates and compared with the log-rank test. Hazard ratio (HR) and 95% confidence interval (CI) were calculated using Cox proportional hazard models. The proportional hazards assumptions of the HRs were graphically inspected in the “log minus log” plot in the Cox proportional hazards models and were tested by Schoenfeld residuals. Propensity-score matched analysis was also performed to reduce selection bias. The covariate balance after propensity-score matching was assessed by calculating absolute standardized mean differences. Standardized mean differences after propensity-score matching were within ± 10% across all matched covariates with variance ratios near 1.0, suggesting achievement of balance between the LMCAD non-culprit group and the no LMCAD group. Stratified Cox proportional hazard models were used to compare the outcomes of the matched groups. All tests were two-tailed, and *p* values < 0.05 were considered statistically significant. Statistical analyses were performed using SPSS version 25 for Windows (SPSS Inc) and R version 3.6.0 (R Foundation for Statistical Computing).

## RESULTS

### Baseline clinical characteristics

Among the 429 patients enrolled in this study, 43 were identified with residual non-culprit LMCAD after index PCI (10.1%, LMCAD non-culprit group) and the remaining 386 (89.9%) comprised the no LMCAD group. The mean age and body mass index of the study population were 68.1 ± 12.1 years and 23.6 ± 3.3, respectively. The incidence of diabetes mellitus was higher in the LMCAD non-culprit group than in the no LMCAD group (*p* = 0.029), but the rate of current smokers was lower in the LMCAD non-culprit group compared to the no LMCAD group (*p* = 0.026). There were no significant differences in type of AMI, left ventricular ejection fraction (LVEF), initial blood pressure, laboratory findings, and emergent in-hospital management between the two groups (**Table 1**). Angiographic and procedural characteristics are presented in Table 2. There was no significant difference in angiographic findings including culprit lesion location or pre- and post-thrombolysis in myocardial infarction (TIMI) flow at the culprit lesion. However, number of diseased coronary vessels (*p* = 0.001), number of stenotic lesions (*p* = 0.001), and synergy between PCI with Taxus and cardiac surgery (SYNTAX) scores before PCI (*p* < 0.001) were significantly different between the two groups. In the procedural characteristics, number of stents used was higher (*p* = 0.042) and thrombus aspiration was performed less frequently (*p* = 0.038) in the LMCAD non-culprit group than in the no LMCAD group (**Table 2**).

**Table 1.**
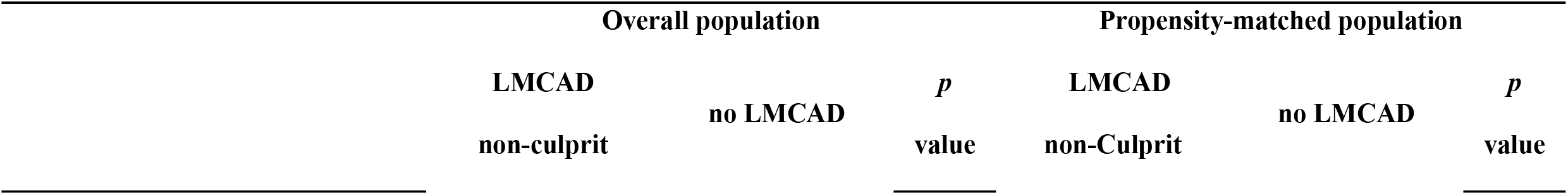
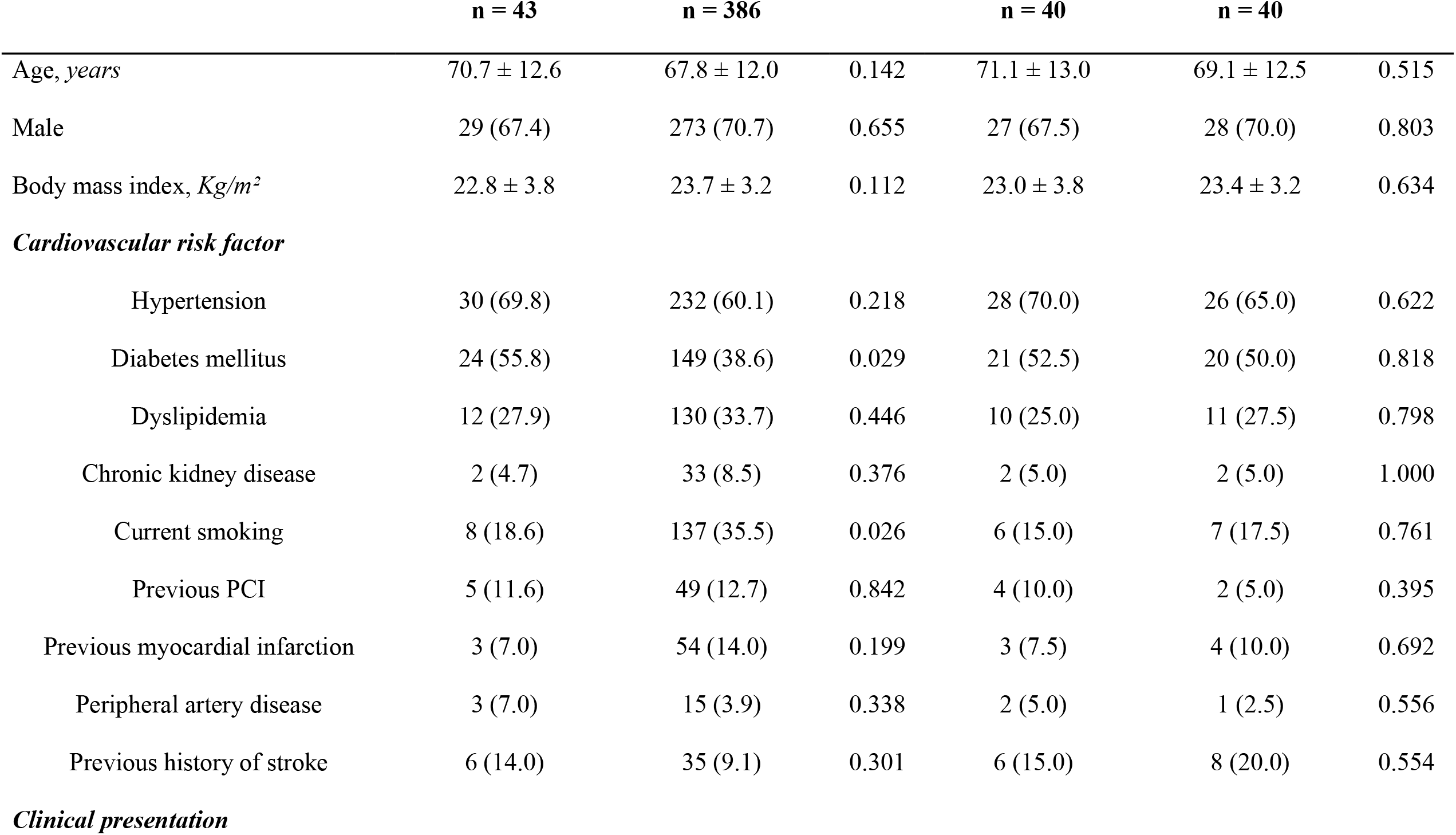
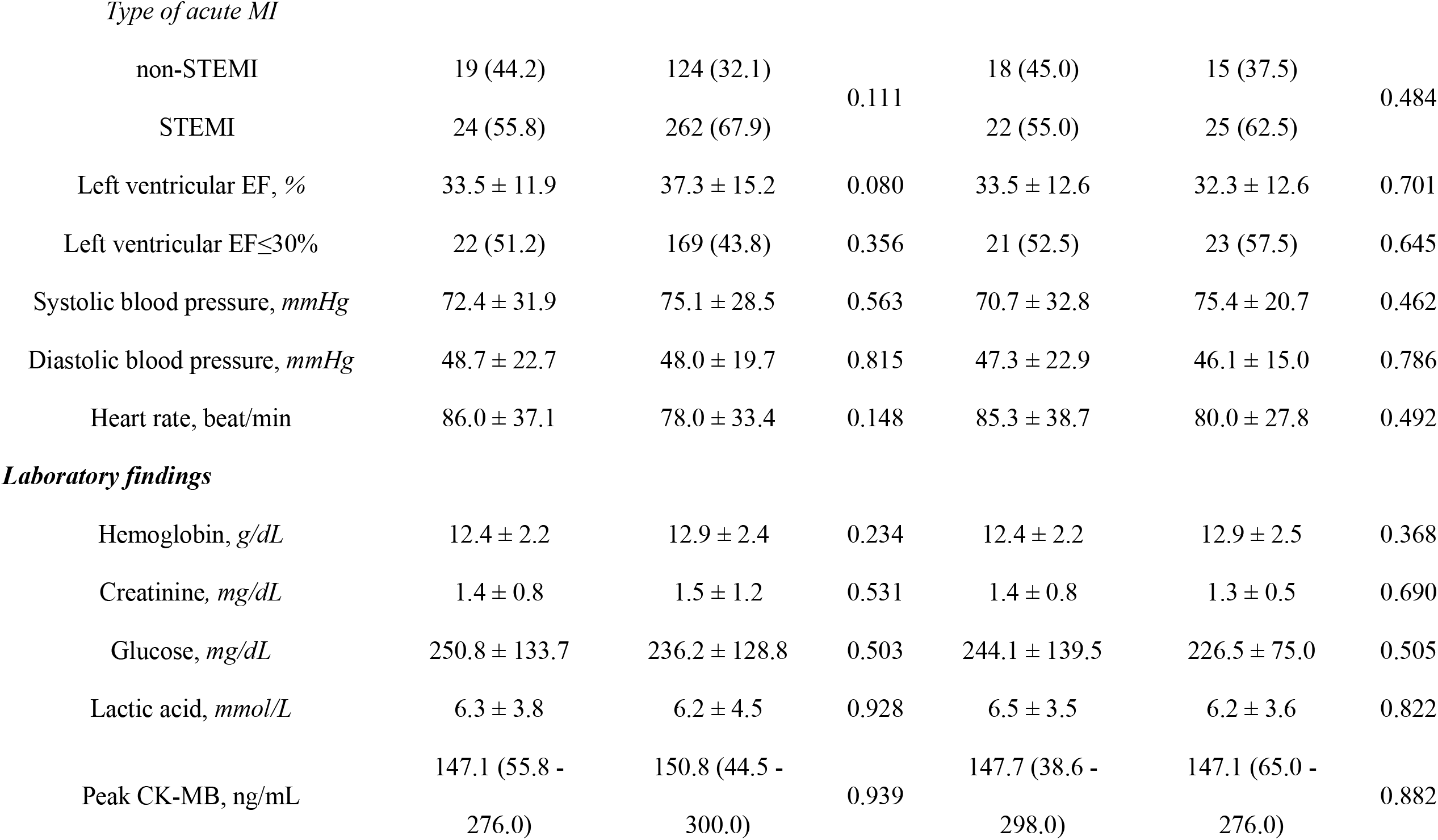
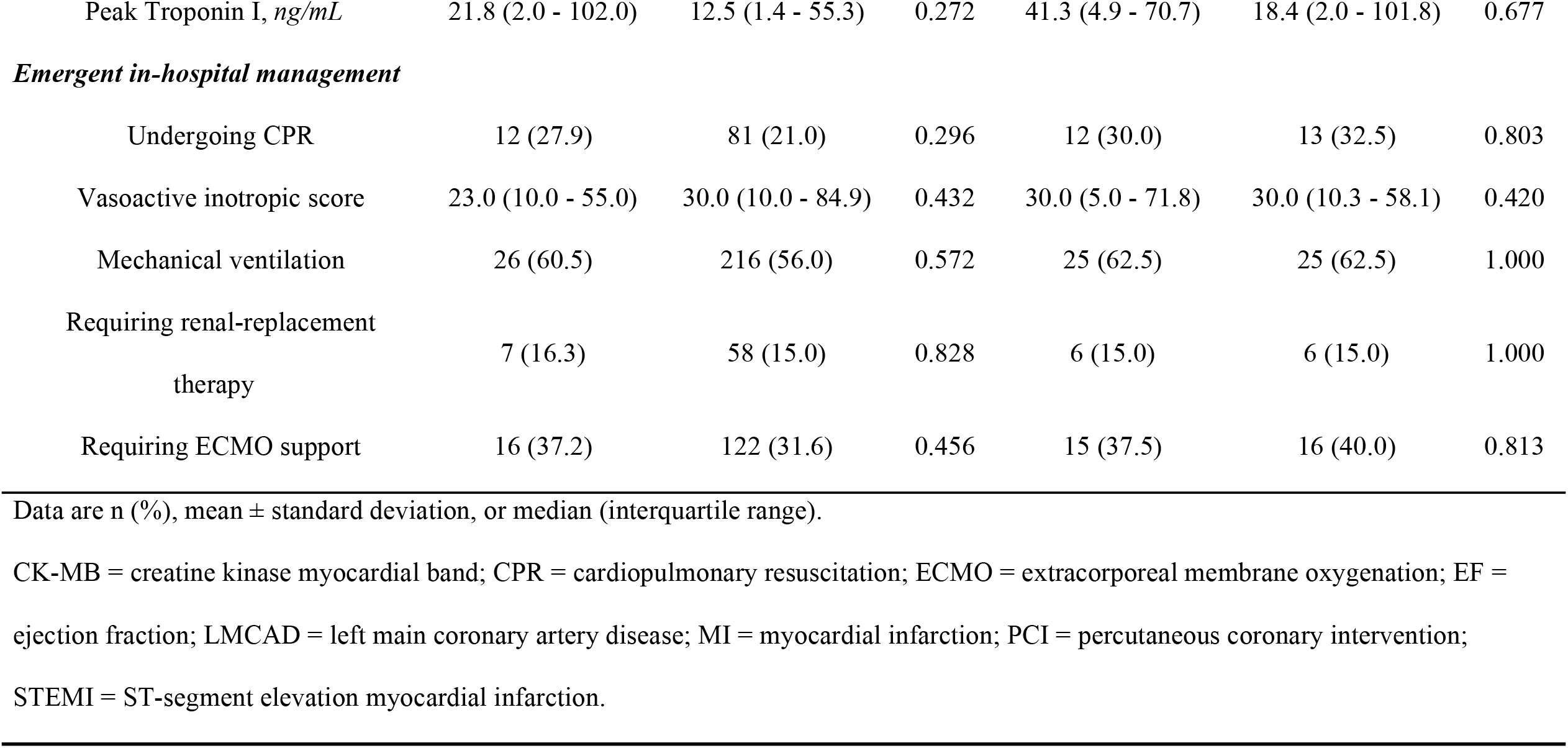
Baseline clinical characteristics and In-hospital management.

| Overall population |  |  | Propensity-matched population |  |  |
| --- | --- | --- | --- | --- | --- |
| LMCAD<br>non-culprit | no LMCAD | *p*<br>value | LMCAD<br>non-Culprit | no LMCAD | *p*<br>value |

|  | **n = 43** | **n = 386** |  | **n = 40** | **n = 40** |  |
| --- | --- | --- | --- | --- | --- | --- |
| Age, years | 70.7 ± 12.6 | 67.8 ± 12.0 | 0.142 | 71.1 ± 13.0 | 69.1 ± 12.5 | 0.515 |
| Male | 29 (67.4) | 273 (70.7) | 0.655 | 27 (67.5) | 28 (70.0) | 0.803 |
| Body mass index, Kg/m<sup>2</sup> | 22.8 ± 3.8 | 23.7 ± 3.2 | 0.112 | 23.0 ± 3.8 | 23.4 ± 3.2 | 0.634 |
| ***Cardiovascular risk factor*** |  |  |  |  |  |  |
| Hypertension | 30 (69.8) | 232 (60.1) | 0.218 | 28 (70.0) | 26 (65.0) | 0.622 |
| Diabetes mellitus | 24 (55.8) | 149 (38.6) | 0.029 | 21 (52.5) | 20 (50.0) | 0.818 |
| Dyslipidemia | 12 (27.9) | 130 (33.7) | 0.446 | 10 (25.0) | 11 (27.5) | 0.798 |
| Chronic kidney disease | 2 (4.7) | 33 (8.5) | 0.376 | 2 (5.0) | 2 (5.0) | 1.000 |
| Current smoking | 8 (18.6) | 137 (35.5) | 0.026 | 6 (15.0) | 7 (17.5) | 0.761 |
| Previous PCI | 5 (11.6) | 49 (12.7) | 0.842 | 4 (10.0) | 2 (5.0) | 0.395 |
| Previous myocardial infarction | 3 (7.0) | 54 (14.0) | 0.199 | 3 (7.5) | 4 (10.0) | 0.692 |
| Peripheral artery disease | 3 (7.0) | 15 (3.9) | 0.338 | 2 (5.0) | 1 (2.5) | 0.556 |
| Previous history of stroke | 6 (14.0) | 35 (9.1) | 0.301 | 6 (15.0) | 8 (20.0) | 0.554 |

***Clinical presentation***
|  |  |  |  |  |  |  |
| --- | --- | --- | --- | --- | --- | --- |
| *Type of acute MI* |  |  |  |  |  |  |
| non-STEMI | 19 (44.2) | 124 (32.1) | 0.111 | 18 (45.0) | 15 (37.5) | 0.484 |
| STEMI | 24 (55.8) | 262 (67.9) |  | 22 (55.0) | 25 (62.5) |  |
| Left ventricular EF, % | 33.5 ± 11.9 | 37.3 ± 15.2 | 0.080 | 33.5 ± 12.6 | 32.3 ± 12.6 | 0.701 |
| Left ventricular EF≤30% | 22 (51.2) | 169 (43.8) | 0.356 | 21 (52.5) | 23 (57.5) | 0.645 |
| Systolic blood pressure, mmHg | 72.4 ± 31.9 | 75.1 ± 28.5 | 0.563 | 70.7 ± 32.8 | 75.4 ± 20.7 | 0.462 |
| Diastolic blood pressure, mmHg | 48.7 ± 22.7 | 48.0 ± 19.7 | 0.815 | 47.3 ± 22.9 | 46.1 ± 15.0 | 0.786 |
| Heart rate, beat/min | 86.0 ± 37.1 | 78.0 ± 33.4 | 0.148 | 85.3 ± 38.7 | 80.0 ± 27.8 | 0.492 |
| *Laboratory findings* |  |  |  |  |  |  |
| Hemoglobin, g/dL | 12.4 ± 2.2 | 12.9 ± 2.4 | 0.234 | 12.4 ± 2.2 | 12.9 ± 2.5 | 0.368 |
| Creatinine, mg/dL | 1.4 ± 0.8 | 1.5 ± 1.2 | 0.531 | 1.4 ± 0.8 | 1.3 ± 0.5 | 0.690 |
| Glucose, mg/dL | 250.8 ± 133.7 | 236.2 ± 128.8 | 0.503 | 244.1 ± 139.5 | 226.5 ± 75.0 | 0.505 |
| Lactic acid, mmol/L | 6.3 ± 3.8 | 6.2 ± 4.5 | 0.928 | 6.5 ± 3.5 | 6.2 ± 3.6 | 0.822 |
| Peak CK-MB, ng/mL | 147.1 (55.8 -<br>276.0) | 150.8 (44.5 -<br>300.0) | 0.939 | 147.7 (38.6 -<br>298.0) | 147.1 (65.0 -<br>276.0) | 0.882 |
| Peak Troponin I, *ng/mL* | 21.8 (2.0 - 102.0) | 12.5 (1.4 - 55.3) | 0.272 | 41.3 (4.9 - 70.7) | 18.4 (2.0 - 101.8) | 0.677 |
| ***Emergent in-hospital management*** |  |  |  |  |  |  |
| Undergoing CPR | 12 (27.9) | 81 (21.0) | 0.296 | 12 (30.0) | 13 (32.5) | 0.803 |
| Vasoactive inotropic score | 23.0 (10.0 - 55.0) | 30.0 (10.0 - 84.9) | 0.432 | 30.0 (5.0 - 71.8) | 30.0 (10.3 - 58.1) | 0.420 |
| Mechanical ventilation | 26 (60.5) | 216 (56.0) | 0.572 | 25 (62.5) | 25 (62.5) | 1.000 |
| Requiring renal-replacement<br>therapy | 7 (16.3) | 58 (15.0) | 0.828 | 6 (15.0) | 6 (15.0) | 1.000 |
| Requiring ECMO support | 16 (37.2) | 122 (31.6) | 0.456 | 15 (37.5) | 16 (40.0) | 0.813 |

Data are n (%), mean ± standard deviation, or median (interquartile range).
CK-MB = creatine kinase myocardial band; CPR = cardiopulmonary resuscitation; ECMO = extracorporeal membrane oxygenation; EF = ejection fraction; LMCAD = left main coronary artery disease; MI = myocardial infarction; PCI = percutaneous coronary intervention; STEMI = ST-segment elevation myocardial infarction.

**Table 2.**
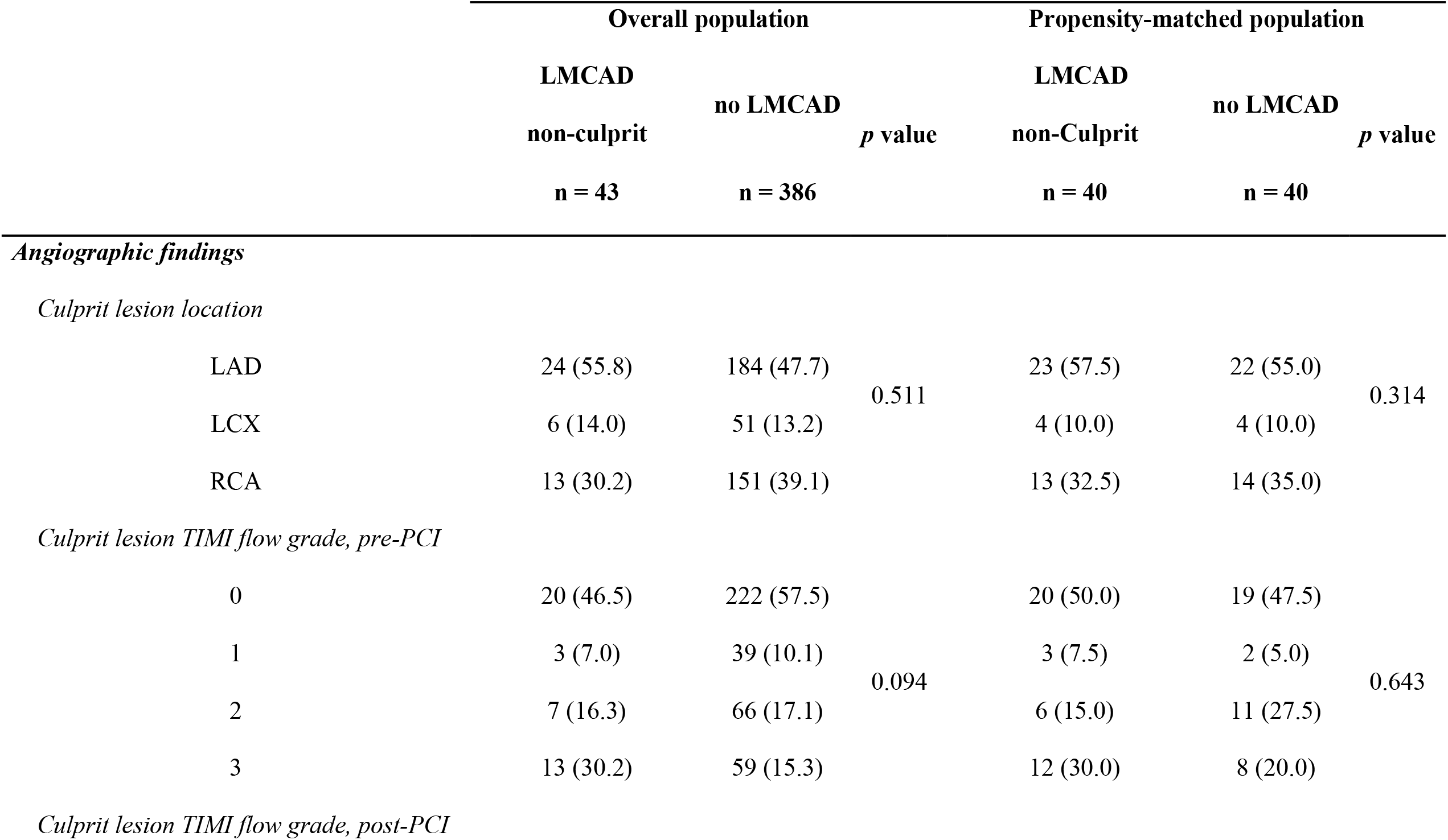
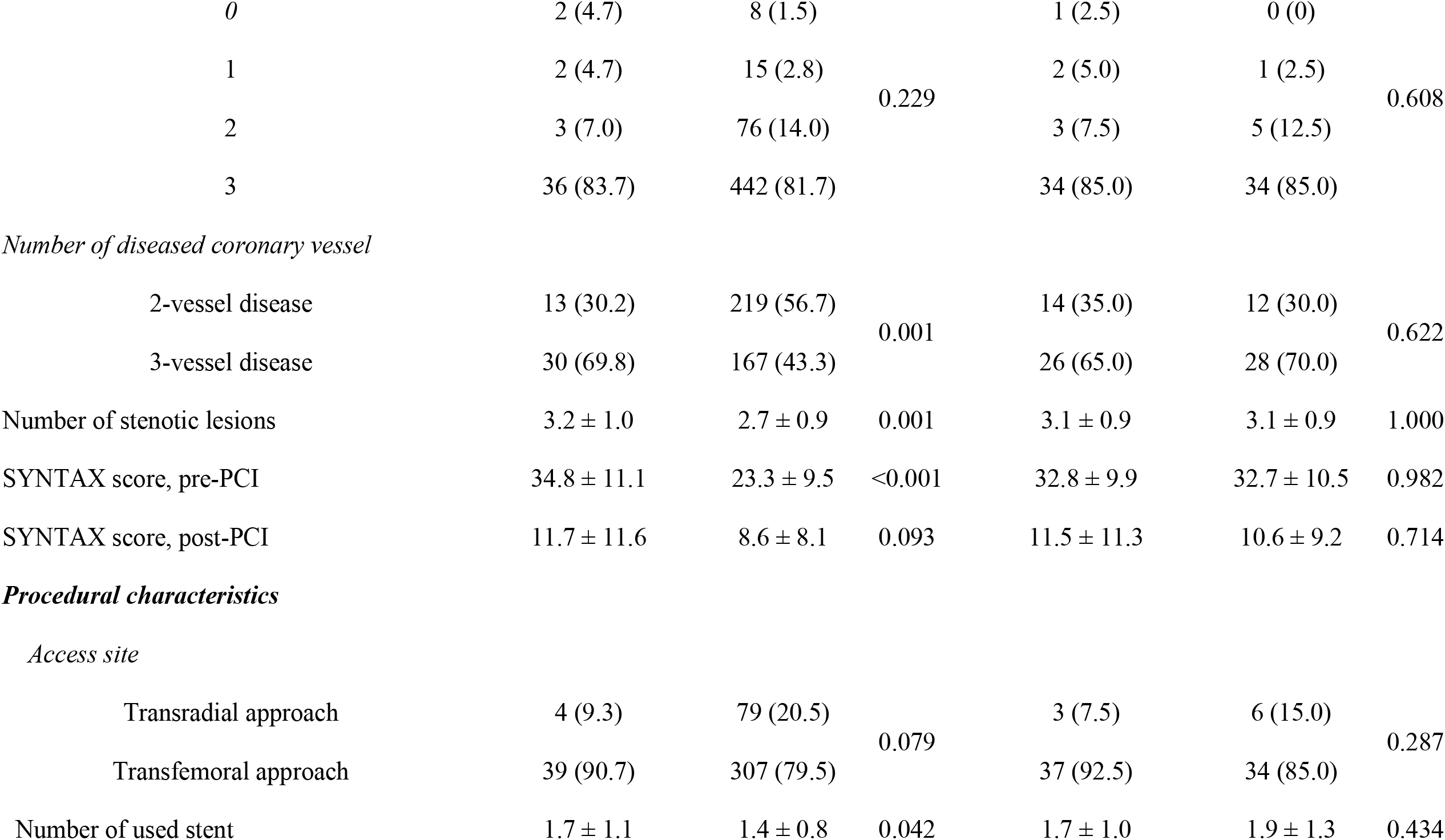
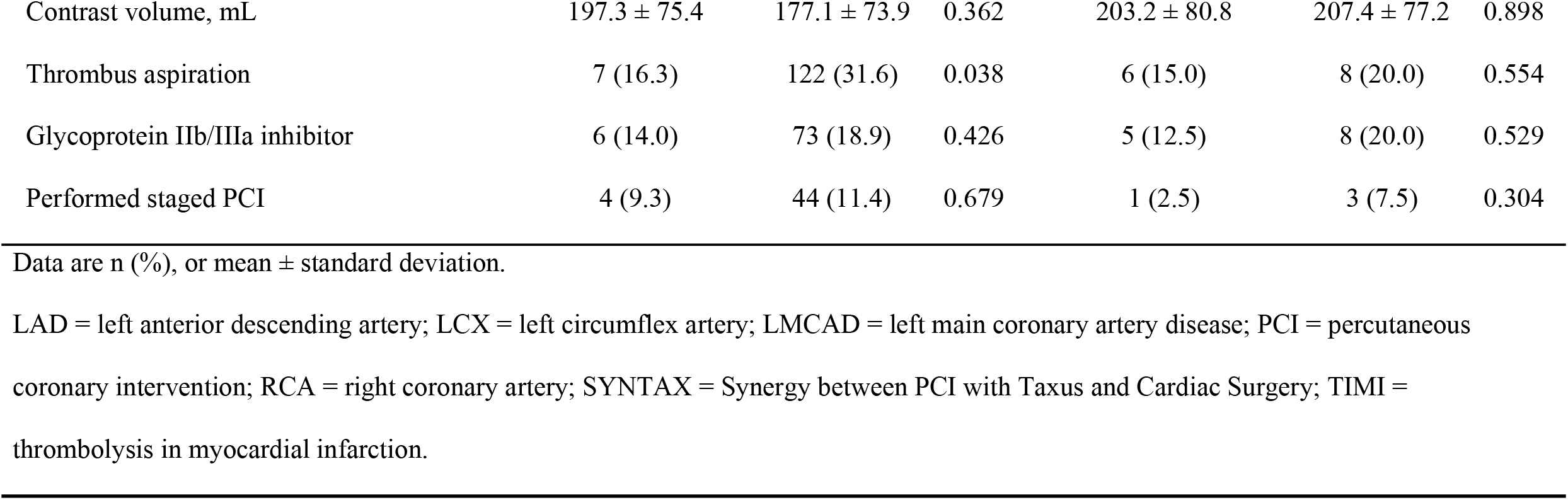
Angiographic and Procedural characteristics.

|  |  | Overall population |  |  | Propensity-matched population |  |  |
| --- | --- | --- | --- | --- | --- | --- | --- |
|  |  | LMCAD |  |  | LMCAD |  |  |
|  |  | non-culprit | no LMCAD | *p* value | non-Culprit | no LMCAD | *p* value |
|  |  | n = 43 | n = 386 |  | n = 40 | n = 40 |  |
| *Angiographic findings* |  |  |  |  |  |  |  |
| *Culprit lesion location* |  |  |  |  |  |  |  |
|  | LAD | 24 (55.8) | 184 (47.7) | 0.511 | 23 (57.5) | 22 (55.0) | 0.314 |
|  | LCX | 6 (14.0) | 51 (13.2) |  | 4 (10.0) | 4 (10.0) |  |
|  | RCA | 13 (30.2) | 151 (39.1) |  | 13 (32.5) | 14 (35.0) |  |
| *Culprit lesion TIMI flow grade, pre-PCI* |  |  |  |  |  |  |  |
|  | 0 | 20 (46.5) | 222 (57.5) | 0.094 | 20 (50.0) | 19 (47.5) | 0.643 |
|  | 1 | 3 (7.0) | 39 (10.1) |  | 3 (7.5) | 2 (5.0) |  |
|  | 2 | 7 (16.3) | 66 (17.1) |  | 6 (15.0) | 11 (27.5) |  |
|  | 3 | 13 (30.2) | 59 (15.3) |  | 12 (30.0) | 8 (20.0) |  |
| *Culprit lesion TIMI flow grade, post-PCI* |  |  |  |  |  |  |  |

|  |  |  |  |  |  |  |
| --- | --- | --- | --- | --- | --- | --- |
| 0 | 2 (4.7) | 8 (1.5) |  | 1 (2.5) | 0 (0) |  |
| 1 | 2 (4.7) | 15 (2.8) | 0.229 | 2 (5.0) | 1 (2.5) | 0.608 |
| 2 | 3 (7.0) | 76 (14.0) |  | 3 (7.5) | 5 (12.5) |  |
| 3 | 36 (83.7) | 442 (81.7) |  | 34 (85.0) | 34 (85.0) |  |
| *Number of diseased coronary vessel* |  |  |  |  |  |  |
| 2-vessel disease | 13 (30.2) | 219 (56.7) | 0.001 | 14 (35.0) | 12 (30.0) | 0.622 |
| 3-vessel disease | 30 (69.8) | 167 (43.3) |  | 26 (65.0) | 28 (70.0) |  |
| Number of stenotic lesions | 3.2 ± 1.0 | 2.7 ± 0.9 | 0.001 | 3.1 ± 0.9 | 3.1 ± 0.9 | 1.000 |
| SYNTAX score, pre-PCI | 34.8 ± 11.1 | 23.3 ± 9.5 | <0.001 | 32.8 ± 9.9 | 32.7 ± 10.5 | 0.982 |
| SYNTAX score, post-PCI | 11.7 ± 11.6 | 8.6 ± 8.1 | 0.093 | 11.5 ± 11.3 | 10.6 ± 9.2 | 0.714 |
| *Procedural characteristics* |  |  |  |  |  |  |
| *Access site* |  |  |  |  |  |  |
| Transradial approach | 4 (9.3) | 79 (20.5) | 0.079 | 3 (7.5) | 6 (15.0) | 0.287 |
| Transfemoral approach | 39 (90.7) | 307 (79.5) |  | 37 (92.5) | 34 (85.0) |  |
| Number of used stent | 1.7 ± 1.1 | 1.4 ± 0.8 | 0.042 | 1.7 ± 1.0 | 1.9 ± 1.3 | 0.434 |
| Contrast volume, mL | 197.3 ± 75.4 | 177.1 ± 73.9 | 0.362 | 203.2 ± 80.8 | 207.4 ± 77.2 | 0.898 |
| Thrombus aspiration | 7 (16.3) | 122 (31.6) | 0.038 | 6 (15.0) | 8 (20.0) | 0.554 |
| Glycoprotein IIb/IIIa inhibitor | 6 (14.0) | 73 (18.9) | 0.426 | 5 (12.5) | 8 (20.0) | 0.529 |
| Performed staged PCI | 4 (9.3) | 44 (11.4) | 0.679 | 1 (2.5) | 3 (7.5) | 0.304 |

Data are n (%), or mean ± standard deviation.
LAD = left anterior descending artery; LCX = left circumflex artery; LMCAD = left main coronary artery disease; PCI = percutaneous coronary intervention; RCA = right coronary artery; SYNTAX = Synergy between PCI with Taxus and Cardiac Surgery; TIMI = thrombolysis in myocardial infarction.

### Clinical outcomes

#### Overall population

Of the study population, 130 all-cause deaths occurred during the initial 30 days after PCI for AMI with CS; 30-day mortality was not significantly different between the LMCAD non-culprit and the no LMCAD groups (16 patients, 37.2% in the LMCAD non-culprit group vs. 114 patients, 29.5% in the no LMCAD group, adjusted HR 1.11 95% CI 0.65 - 1.91, *p* = 0.703) (**Supplemental Table 1**). By 12 months after the index procedure, the primary outcome had occurred in 17 patients (39.5%) in the LMCAD non-culprit group and 151 patients (39.1%) in the no LMCAD group (adjusted HR 0.94, 95% CI 0.56 - 1.58; *p* = 0.817). There were no significant differences in the individual components of the primary outcome (cardiac death 34.9% in the LMCAD non-culprit group vs. 33.7% in the no LMCAD group, adjusted HR 0.95, 95% CI 0.55 - 1.65, *p* = 0.868; myocardial infarction 4.7% vs.2.3%, adjusted HR 1.29, 95% CI 0.25 - 6.58, *p* = 0.762; repeat revascularization 2.3% vs. 4.4%, adjusted HR 0.45, 95% CI 0.06 - 3.73, *p* = 0.461), all-cause death (46.5% vs. 41.5%, adjusted HR 1.03, 95% CI 0.64 - 1.66, *p* = 0.904), and re-hospitalization due to heart failure (4.7% vs. 6.2%, adjusted HR 0.63, 95% CI 0.15 - 2.95, *p* = 0.599) at 12 months (**Figure 2** and **Table 3**).

**Table 3.**
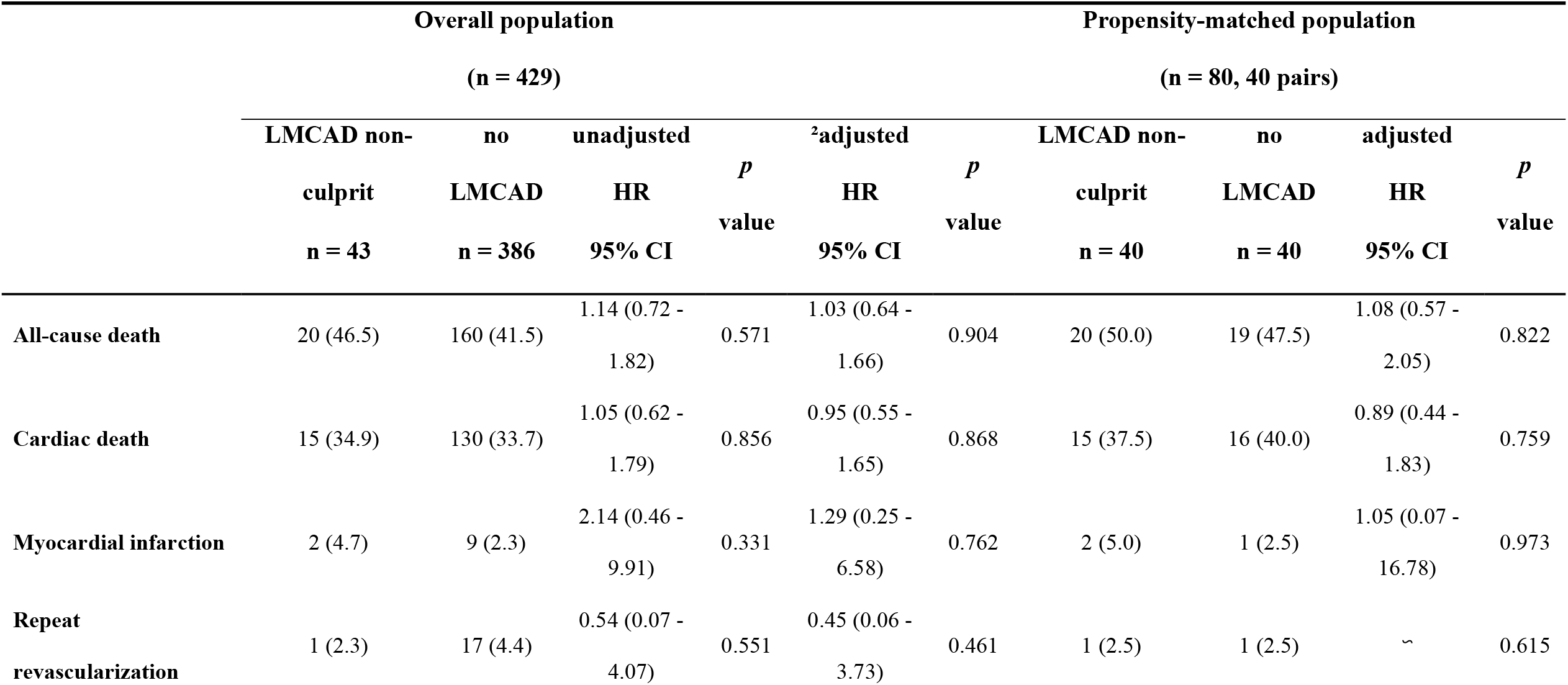
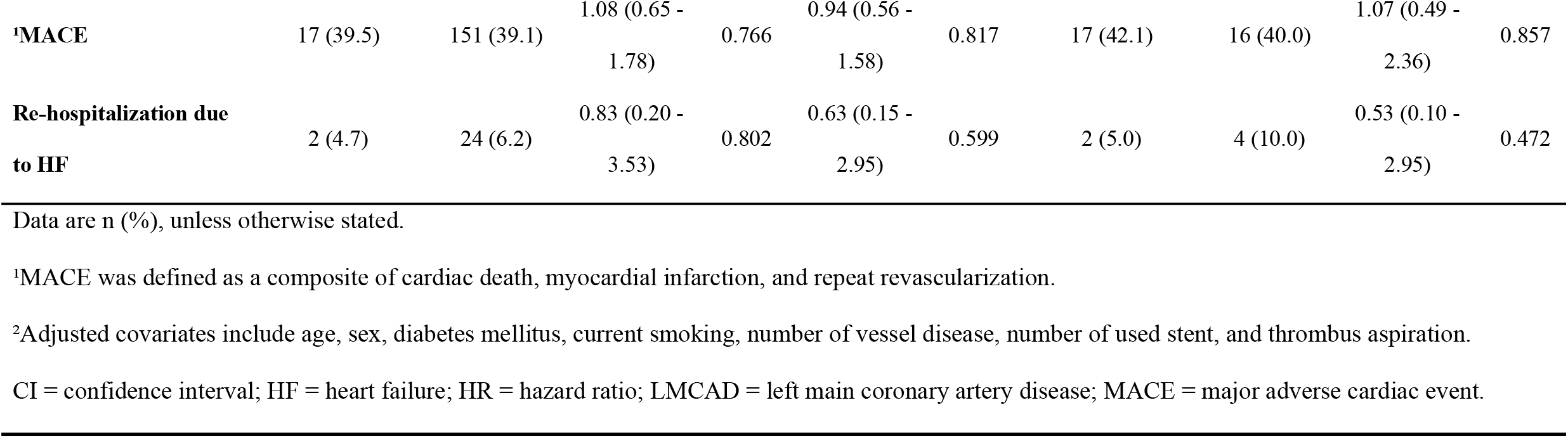
Clinical Outcomes during 12-month Follow-Up.

|  | Overall population |  |  |  |  |  | Propensity-matched population |  |  |  |
| --- | --- | --- | --- | --- | --- | --- | --- | --- | --- | --- |
|  | (n = 429) |  |  |  |  |  | (n = 80, 40 pairs) |  |  |  |
|  | LMCAD non- | no | unadjusted |  | <sup>2</sup>adjusted |  | LMCAD non- | no | adjusted |  |
|  | culprit | LMCAD | HR | *p* | HR | *p* | culprit | LMCAD | HR | *p* |
|  | n = 43 | n = 386 | 95% CI | value | 95% CI | value | n = 40 | n = 40 | 95% CI | value |
| All-cause death | 20 (46.5) | 160 (41.5) | 1.14 (0.72 - 1.82) | 0.571 | 1.03 (0.64 - 1.66) | 0.904 | 20 (50.0) | 19 (47.5) | 1.08 (0.57 - 2.05) | 0.822 |
| Cardiac death | 15 (34.9) | 130 (33.7) | 1.05 (0.62 - 1.79) | 0.856 | 0.95 (0.55 - 1.65) | 0.868 | 15 (37.5) | 16 (40.0) | 0.89 (0.44 - 1.83) | 0.759 |
| Myocardial infarction | 2 (4.7) | 9 (2.3) | 2.14 (0.46 - 9.91) | 0.331 | 1.29 (0.25 - 6.58) | 0.762 | 2 (5.0) | 1 (2.5) | 1.05 (0.07 - 16.78) | 0.973 |
| Repeat revascularization | 1 (2.3) | 17 (4.4) | 0.54 (0.07 - 4.07) | 0.551 | 0.45 (0.06 - 3.73) | 0.461 | 1 (2.5) | 1 (2.5) | ~ | 0.615 |
| **<sup>1</sup>MACE** | 17 (39.5) | 151 (39.1) | 1.08 (0.65 - 1.78) | 0.766 | 0.94 (0.56 - 1.58) | 0.817 | 17 (42.1) | 16 (40.0) | 1.07 (0.49 - 2.36) | 0.857 |
| **Re-hospitalization due to HF** | 2 (4.7) | 24 (6.2) | 0.83 (0.20 - 3.53) | 0.802 | 0.63 (0.15 - 2.95) | 0.599 | 2 (5.0) | 4 (10.0) | 0.53 (0.10 - 2.95) | 0.472 |

Data are n (%), unless otherwise stated.
<sup>2</sup>Adjusted covariates include age, sex, diabetes mellitus, current smoking, number of vessel disease, number of used stent, and thrombus aspiration.
CI = confidence interval; HF = heart failure; HR = hazard ratio; LMCAD = left main coronary artery disease; MACE = major adverse cardiac event.

**Figure 2.**
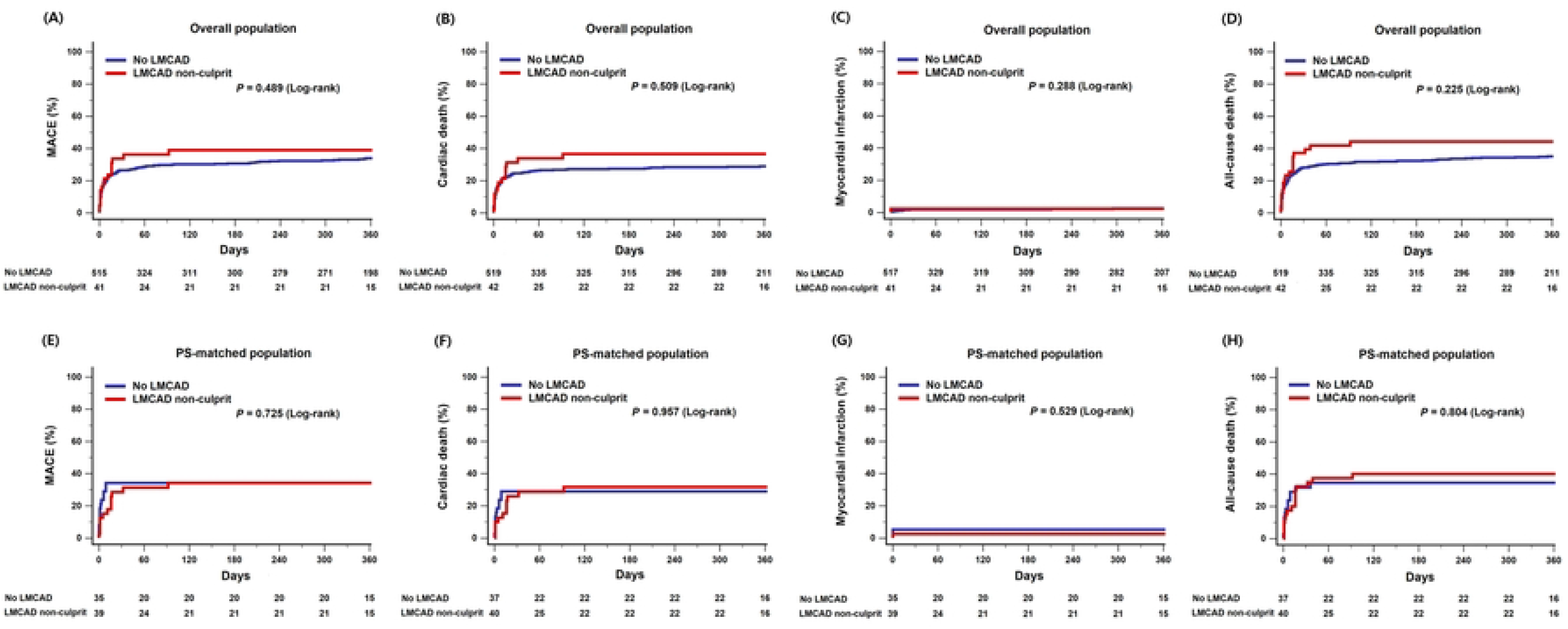
Time-to-event Kaplan-Meier survival curves of clinical outcome according to presence of non-culprit LMCAD. (A) Kaplan-Meier curves for major adverse cardiac event (MACE) and (E) MACE after propensity-matched adjustment. (B) Kaplan-Meier curves for cardiac death and (F) cardiac death after propensity-matched adjustment. (C) Kaplan-Meier curves for myocardial infarction and (E) myocardial infarction after propensity-matched adjustment. (D) Kaplan-Meier curves for all-cause death and (F) all-cause death after propensity-matched adjustment. MACE was defined as a composite of cardiac death, myocardial infarction, and repeat revascularization. LMCAD = left main coronary artery disease; MACE = major adverse cardiac event.

#### Propensity-matched population

After performing propensity score matching, a total of 40 pairs was generated. There were no significant differences in baseline clinical or angiographic characteristics for the propensity score-matched subjects (Tables 1 and 2). A total of 33 MACEs occurred during follow-up in matched patients, and there was no significant difference in the incidence of MACEs at 12 months (matched HR 1.07, 95% CI 0.49 - 2.36; *p* = 0.857) between the LMCAD non-culprit and the no LMCAD groups. The risk of cardiac death (matched HR 0.89, 95% CI 0.44 - 1.83; *p* = 0.759), all-cause death (matched HR 1.08, 95% CI 0.57 - 2.05; *p* = 0.822), MI (5.0% vs 2.5%; *p* = 0.973), repeat revascularization (2.5% vs 2.5%, *p* = 0.615), and re-hospitalization due to heart failure (matched HR 0.53, 95% CI 0.10 - 2.95; *p* = 0.472) were also similar between the two groups (**Figure 2** and **Table 3**).

### Subgroup analysis

To investigate the association between presence of non-culprit LMCAD and MACE after PCI for AMI with CS in various situations, we performed subgroup analyses. The prognostic effect of residual non-culprit LMCAD did not differ significantly across subgroups regardless of age (≥ 65 years vs. < 65 years), body mass index (≥ 25.0 vs. < 25.0), sex, presence of diabetes mellitus, type of AMI (STEMI vs. non-STEMI), LVEF < 30%, extracorporeal membrane oxygenation (ECMO) support, initial serum lactate (< 8.0 vs ≥ 8.0), or vasoactive inotropic score (< 84.0 vs. ≥ 84.0) (**Figure 3**).

**Figure 3.**
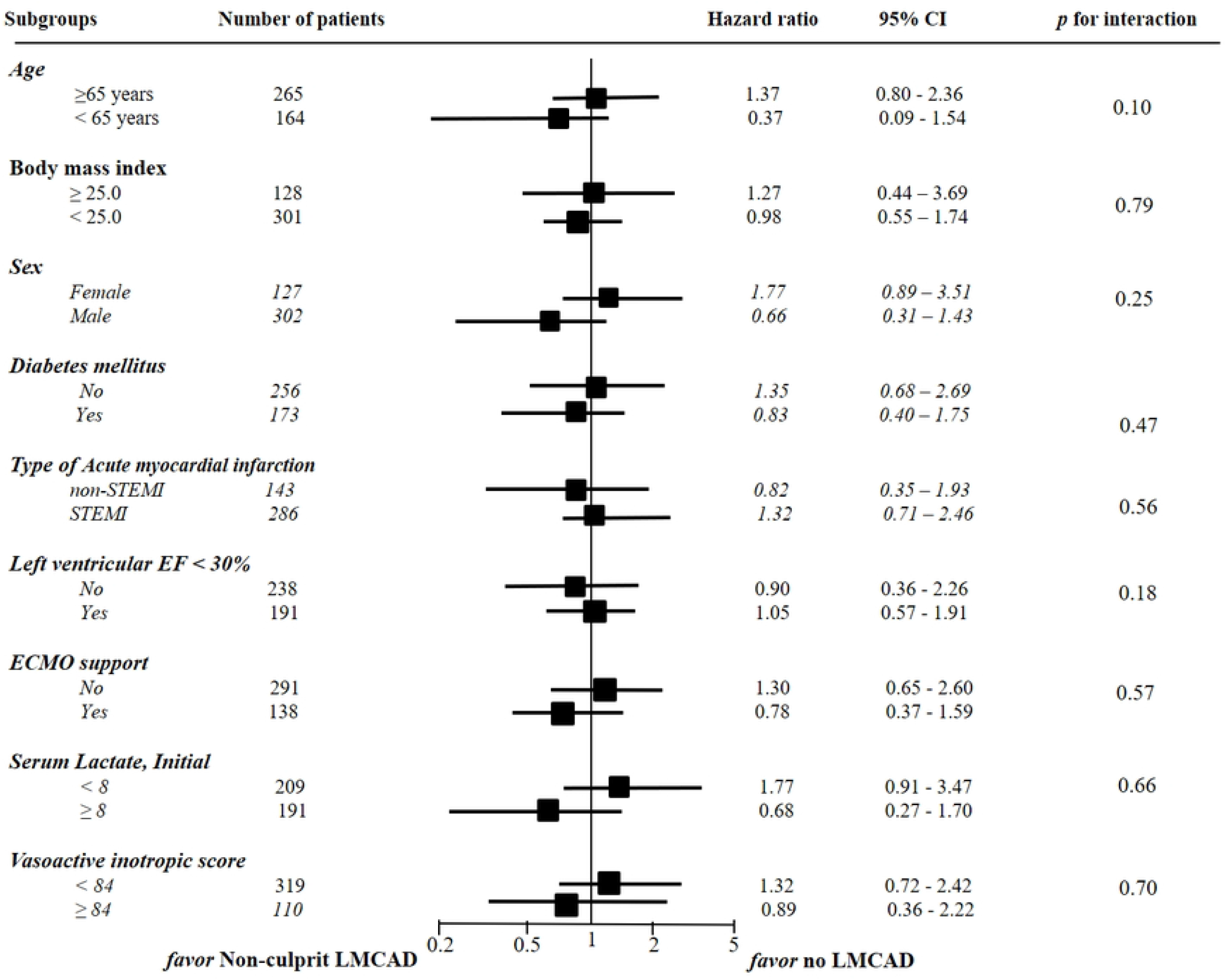
Comparative unadjusted subgroup hazard ratios for primary outcome between non-culprit LMCAD and no LMCAD groups. MACE was defined as a composite of cardiac death, myocardial infarction, and repeat revascularization. CI = confidence interval; ECMO = extracorporeal membrane oxygenation; EF = ejection fraction; LMCAD = left main coronary artery disease; MACE = major adverse cardiac event; STEMI = ST-segment elevation myocardial infarction

## DISCUSSION

This study investigated the clinical impact of non-culprit LMCAD on 12-month clinical outcomes in patients treated with PCI for AMI complicated by CS using a dedicated, large-scale, multicenter real-world CS registry. The main study finding was that there was no significant difference in risk of a composite of cardiac death, myocardial infarction, or repeat revascularization during 12 months between the LMCAD non-culprit group and the no LMCAD group. This was consistent across subgroups by use of ECMO support and a variety of other clinical factors. To the best of our knowledge, this is the first study specifically concerned with the prognostic effect of residual non-culprit LMCAD in patients undergoing emergent PCI for AMI complicated by CS.

PCI for LMCAD using a drug-eluting stent could be safe and effective in stable coronary artery disease (CAD). However, particularly in the presence of shock, the interventional treatment of LMCAD, either culprit- or non-culprit lesion, is always a lot of concern. The reason for this is the need for multiple stents and the complex of PCI, which is associated with acute stent thrombosis or chronic target lesion revascularization [9]. Moreover, despite the findings of many previous studies, it has remained unclear whether residual non-culprit LMCAD are translated into adverse clinical outcomes [9-11]. Also, the optimal treatment strategy for non-culprit LMCAD in AMI with CS is unknown. Approximately 50% of patients undergoing PCI for AMI with CS have a significant multi-vessel CAD including LMCAD [1,10]. Because AMI with CS is an emergent status that represents high thrombus burden with the possibility of undersized stenting in PCI, the comparative performance of coronary stents in such a critical shock setting is an area of substantial uncertainty [12]. Coronary artery bypass grafting could have a better outcome compared to PCI in cases of stable LMCAD, but surgical treatment frequently may not be applicable in cases of AMI complicated by CS [13]. Several randomized controlled trials have demonstrated that CR for significant non-culprit stenoses, either at the time of the index procedure or as a staged procedure, is superior to a culprit-only treatment in reducing the risk for cardiovascular events among patients with multi-vessel CAD [2,14]. Similar previous studies performed in patients with LMCAD also comprised a significant proportion of stable ischemic heart disease patients but always excluded CS patients [15,16]. Maron et al. [3] investigated patients with stable CAD with moderate or severe ischemia. This group did not find any evidence that an initial invasive strategy reduced the risk of ischemic cardiovascular events or death from any cause, but CS patients were also excluded from their study. The optimal strategy to guide revascularization of non-culprit stenosis on LMCAD in patients with AMI and CS remains uncertain after these previous studies and in current guidelines. Moreover, the prognostic impact of residual non-culprit LMCAD after PCI for multi-vessel CAD is virtually unknown in patients with AMI complicated by CS. Therefore, our study addressed the clinical impact of non-culprit LMCAD on 12-month clinical outcomes in patients treated with PCI for AMI complicated by CS. We identified that there was no significant difference in clinical prognosis during the 12 months after the index procedure between CS patients with or without residual non-culprit LMCAD. Thiele et al. showed that culprit-lesion-only PCI had lower 30-day risk of adverse outcome compared to immediate multi-vessel PCI among patients who had multi-vessel CAD and AMI with CS. Despite the short-term nature of this evaluation, the results correspond well with those of our study [10]. In subgroup analysis, the similarity of MACE incidence between the LMCAD non-culprit and the no LMCAD groups was consistent across analysis of a variety of clinical factors. In particular, LVEF, vasoactive inotropic score, and provision of ECMO support were used as clinical variables to evaluate interactions between residual non-culprit LMCAD and CS severity. Previous studies have suggested significant associations among low LVEF, high vasoactive inotropic score, or requirement of ECMO support and adverse clinical outcomes in CS patients [4,17]. During at least 12 months after index PCI, there were no significant interactions between residual non-culprit LMCAD and clinical outcomes according to CS severity in our study.

Interestingly, our patients with non-culprit LMCAD were treated with only medical therapy during the 12-month follow-up period. The one exception was a patient who was treated with repeat PCI at six months because of in-stent restenosis of a previous culprit lesion. Most of the patients with non-culprit LMCAD did not undergo repeat revascularization, and study patients with or without non-culprit LMCAD had similar clinical outcomes. Therefore, we conclude that conservative or optimal medical treatment seems to not be inferior to aggressive or interventional treatment strategy for non-culprit LMCAD within 12 months after culprit PCI in AMI with CS. This conclusion corresponds well with the ISCHEMIA subgroup analysis result that an invasive strategy did not reduce the risk of ischemic cardiovascular events or death from any cause. This subgroup analysis was of cases of two or more vessels or ≥ 50% stenosis from the ostium to proximal vessel on the left anterior descending coronary artery [3].

### Study limitations

Despite the strengths of this study that resulted from the use of a large, dedicated CS registry with minimal exclusion criteria, the study also has several limitations. First, this study was derived from multi-center observational data; unmeasured confounding factors could have influenced the study results. In particular, the choice of revascularization strategy and application of ECMO were at the operator’s discretion, possibly introducing selection bias. Second, although the present registry is the largest to date, the cohort is still relatively small. In addition, the present registry included patients who were treated only with PCI. Some patients may have been treated conservatively, and we did not have any other data about medical treatments. Also, thrombolysis or CABG and all possible clinical outcomes of this type of lesion are not reflected in our results. Third, the lack of significant interaction in certain subgroup analyses may have been due to the limited sample size. Therefore, the current results should be interpreted as hypothesis-generating and should be confirmed in a future, well-designed randomized trial. Fourth, the rate of nonfatal events was low relative to that of death during follow-up. Although we performed active follow-up, periodic site monitoring, and auditing of the source document in each individual center to ensure that all information was properly entered in the electronic case report form, we cannot rule out the possibility of missed events. Finally, the analysis was limited to only 12 months of follow-up. The true difference in prognostic effect of non-culprit LMCAD might not be apparent at 12 months; a longer follow-up duration may be necessary to confirm the clinical impact of non-culprit LMCAD on adverse outcomes in AMI with CS.

## CONCLUSIONS

In patients treated with PCI for AMI complicated by CS, there was no significant difference in the 12-month risk of MACE and secondary outcomes between the LMCAD non-culprit and the no LMCAD groups. The similarity of 12-month MACE between the two groups was consistent across various subgroups. Based on our results, residual non-culprit LMCAD does not seem to influence clinical outcomes for 12 months after the index PCI in patients with AMI complicated by CS. Further investigations in a shock setting are required to confirm this finding.

## Data Availability

All relevant data are within the manuscript and its Supporting Information files.

## ABBREVIATIONS

AMI: acute myocardial infarction
CS: cardiogenic shock
ECMO: extracorporeal membrane oxygenation
LMCAD: left main coronary artery disease
MACE: major adverse cardiac event
PCI: percutaneous coronary intervention
STEMI: ST-segment elevation myocardial infarction

## ACKNOWLEDGMENTS

None

